# Pandemic effect on body composition. Single center analysis of 2.771 cases

**DOI:** 10.1101/2021.03.29.21254558

**Authors:** Fabrício Braga, Roberto Zagury, Cristiane Perroni, Victor Hugo Domecg

**Author notes:** Corresponding author: Fabrício Braga, MSc, Laboratório de Performance Humana, Largo do Ibam, n01 - 2ºfloor - Humaitá, Rio de Janeiro - RJ, Brazil ZIP code: 22271-070.

## Abstract

**Background:** The COVID-19 pandemic has led to a dramatic increase in the levels of sedentary lifestyle and unhealthy dietary habits. A worsening in populational obesity levels and body composition (BC) is strongly awaited but so far not documented.

**Objective:** To compare BC profile measured by bioelectrical impedance analysis (BIA) between pre-pandemic (P1-03/15^th^/2017 to 03/16^th^/2020) and pandemic (P2-3/17^th^/2020 to 3/10^th^/2021) period of time.

**Materials and Methods:** BIA were grouped according to the time it was performed. Two comparisons were done: an independent sample comparison (ISC) and a paired sample comparison (PSC) considering patients with at least one BIA in P1 and P2. Age, height, gender, weight, body mass index (BMI), body fat mass (BFM), free fat mass (FFM), skeletal muscle mass (SMM), percentage of body fat (PBF), visceral fat area (VFA) were compared. Statistical significance level was defined for a p value<0.05.

**Results and Discussion:** A total of 3.358 BIA were performed, and 2.771 and 112 were selected for IS and PS, respectively. In ISC, despite an unchanged weight, BFM, FFM, PBF and VFA increased and SSM decreased on P2(p<0.015 for all). A multivariated linear regression model using PBF as dependent variable showed P2 as an independent predictor (β=0.38 95%CI 0.19 to 0.56). In the PSC, PBF also increased from P1 to P2 (p=0.015). To our knowledge this is the first documentation of worsening BC after pandemic. Health authorities should be alert for this phenomenon and their clinical consequences in the days to come.

## 1. BACKGROUND

The COVID-19 pandemic has led to a dramatic increase in the levels of sedentary lifestyle and also to a worsening in the dietary habits of the population(1, 2). This was a consequence of the necessary social isolation recommended to try to control the spread of the SARS-CoV-2 and induced a huge social, financial, and psychological burden(3). It is well known that both the sitting time and the total weekly volume of training are linked to cardiovascular disease risk and mortality(4, 5). To be confined do not obligatory equates to be sedentary.

It would not be surprising to see an important rise in the number of people with overweight or obesity after the end of this period of imposed home reclusion. But of utmost importance is also the body composition (BC). Not only the weight is a driver of the cardiovascular risk. The percentage of body fat and the area of visceral fat are also important markers of cardiometabolic clinical endpoints(6–8).

As far as we know there are no trials assessing the amount of body fat gained since the beginning of the COVID-19 pandemic in Brazil at a population level. To assess the magnitude of the detrimental effects of this period of worsened lifestyle has a clinical relevance as it may inform public health authorities about one of the downsides of the social isolation that should be kept in mind when one evaluates the risk-to-benefit analysis that precedes the decision regarding this sanitary approach.

The objective of this retrospective analysis is to compare the profile of BC through bioelectrical impedance analysis (BIA) in a large population referred for physical evaluation in an exercise medicine outpatient clinic, before and after SARS-CoV-2 pandemic.

## 2. MATERIALS AND METHODS

BIA carried out between 15^th^ March 2017 and 10^th^ March 2021, were analyzed, all of them using the device Inbody® 770 (Inbody Co., LTD, Seoul, Korea). BIA was performed as a routine measurement before cardiopulmonary exercise test. All individuals were instructed according to the recommendation previously published(9).

Two analysis were done. In the first one, only one measure per individual was taken in account. For those with repeated measures only the first one was considered. For comparison BIA were split in two groups according to period it was performed: Pre-pandemic (between March 15^th^, 2017 and March 16^th^, 2020) and pandemic (between March 17th, 2020 and March 10th, 2021). Age, height, gender, weight (W), body mass index (BMI), body fat mass (BFM), free fat mass (FFM), skeletal muscle mass (SMM), percentage of body fat (PBF), visceral fat area (VFA) were compared between the periods of time.

In the second analysis, we compared BIA for the same individual performed in the pre-pandemic and pandemic time frames. In case of more than a single measurement in one of the periods of time, only the first was taken for analysis. Age, W, BMI, BFM, FFM, SMM, PBF, and VFA were compared between the periods of time.

Because of the exploratory nature of the study no sample size calculation was performed, and an all-comers design was adopted.

Data distribution for each variable was analyzed using Kolmogorov-Smirnov test. Continuous variables were expressed in mean ± SD and were compared by t-student test and by paired t-student test for the first and second analysis, respectively. Categorical variables were expressed as percentage and compared with Chi-squared test. Effect size (ES) was measured by Cohen’s *d*. The qualitative assessment of the ES was interpreted as follow: <0.2: very small; 0.2 to 0.49: small; 0.5 to 0.79: medium, 0.8 to 1.19: large, 1.2 to 1.99 very large and ≥2.0 huge. A linear regression analysis was performed using PBF as dependent variable, and age, gender, W, height, SSM and the period of time as independents variables. Statistical significance level was defined for a p value<0.05.

### Pre-pandemic Pandemic Cohen’s d (95%CI) p value

Statistical analysis was carried out using the Statistical Package for the Social Sciences software (IBM SPSS Statistics for Windows, version 22.0, IBM Corp., Armonk, NY) and Jamovi [The Jamovi project (2021). Jamovi (Version 1.6)].

This study was approved by the Ethics Committee of the Hospital Federal de Bonsucesso under the protocol number 33729120.5.0000.5253 (http://plataformabrasil.saude.gov.br/). All the procedures in this study were in accordance with the 1975 Helsinki Declaration, updated in 2013.

## 3. RESULTS

The selection of participants for the present study is summarized in figure 1.

**Figure 1.**
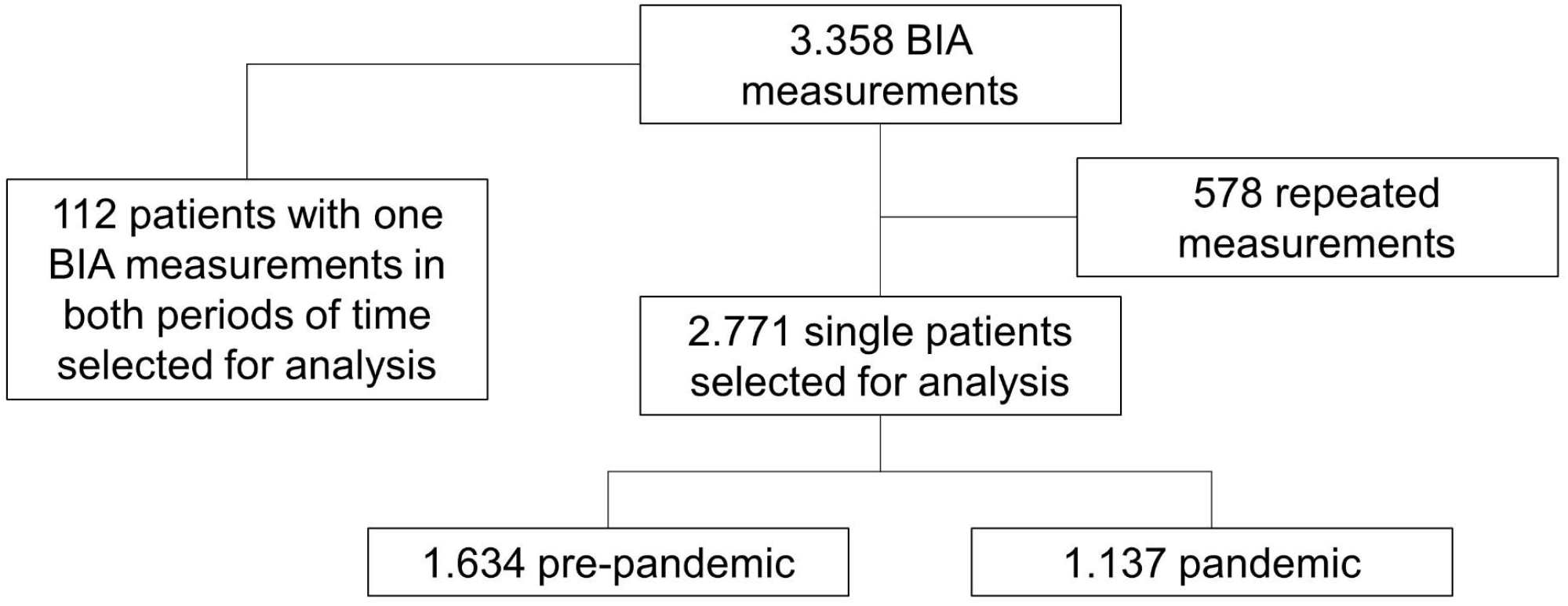
Flowchart of case selection for analysis

For the first analysis, 2.771 patients were included, 1.634 and 1.137 in the pre-pandemic and pandemic time frames, respectively. Table 1 summarizes the univariate analysis.

**Table 1.**
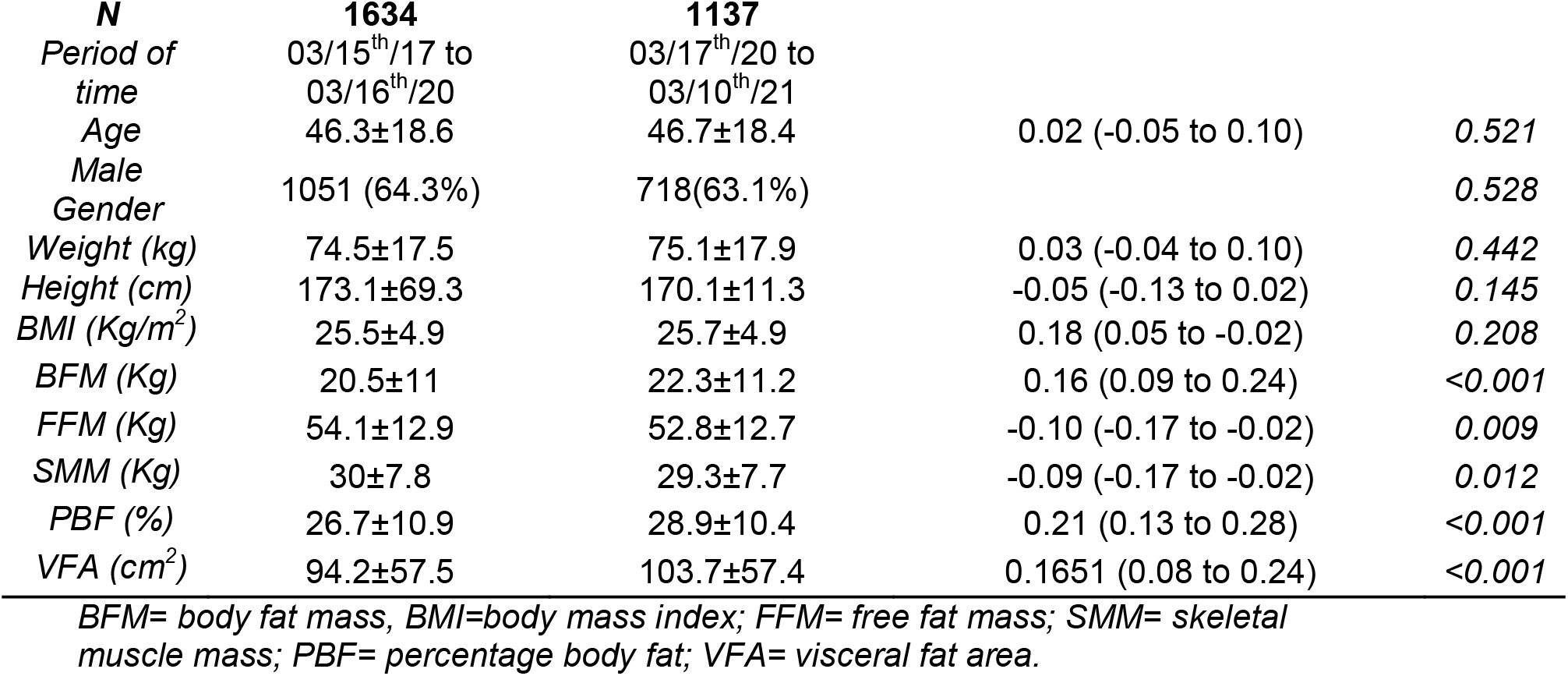
Univariated analysis comparing Pre-pandemic with pandemic time.

| **N** | **1634** | **1137** |  |  |
| --- | --- | --- | --- | --- |
| Period of time | 03/15<sup>th</sup>/17 to 03/16<sup>th</sup>/20 | 03/17<sup>th</sup>/20 to 03/10<sup>th</sup>/21 |  |  |
| Age | 46.3±18.6 | 46.7±18.4 | 0.02 (-0.05 to 0.10) | 0.521 |
| Male Gender | 1051 (64.3%) | 718(63.1%) |  | 0.528 |
| Weight (kg) | 74.5±17.5 | 75.1±17.9 | 0.03 (-0.04 to 0.10) | 0.442 |
| Height (cm) | 173.1±69.3 | 170.1±11.3 | -0.05 (-0.13 to 0.02) | 0.145 |
| BMI (Kg/m<sup>2</sup>) | 25.5±4.9 | 25.7±4.9 | 0.18 (0.05 to -0.02) | 0.208 |
| BFM (Kg) | 20.5±11 | 22.3±11.2 | 0.16 (0.09 to 0.24) | <0.001 |
| FFM (Kg) | 54.1±12.9 | 52.8±12.7 | -0.10 (-0.17 to -0.02) | 0.009 |
| SMM (Kg) | 30±7.8 | 29.3±7.7 | -0.09 (-0.17 to -0.02) | 0.012 |
| PBF (%) | 26.7±10.9 | 28.9±10.4 | 0.21 (0.13 to 0.28) | <0.001 |
| VFA (cm<sup>2</sup>) | 94.2±57.5 | 103.7±57.4 | 0.1651 (0.08 to 0.24) | <0.001 |

BFM= body fat mass, BMI=body mass index; FFM= free fat mass; SMM= skeletal muscle mass; PBF= percentage body fat; VFA= visceral fat area.

Although W did not change, BFM, FFM, PBF and VFA increased and SMM decrease in pandemic time compared to pré-pandemic (figure 2). A small ES was observed for PBF, and a very small ES for the all the others BC variables.

**Figure 2.**
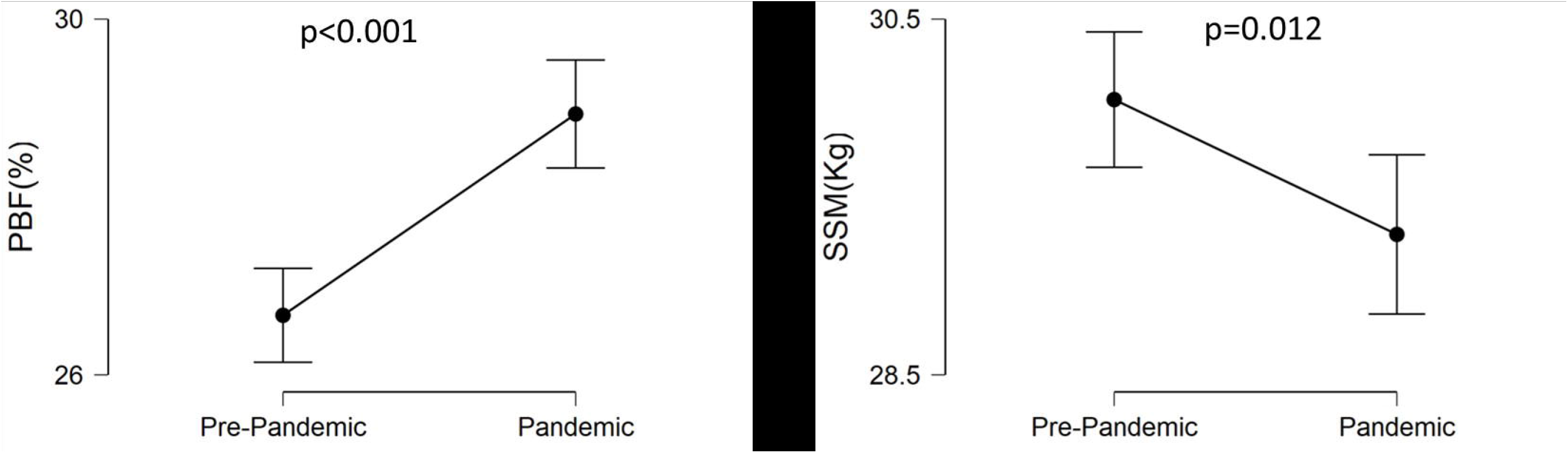
Descriptive plot showing 95% confidence intervals for the mean PBF(A) and SSM(B) in pre-pandemic and pandemic periods of time.

Table 2 shows the multivariate linear regression analysis. After adjustment for previously mentioned variables the pandemic time frame was directly related to PBF (β=0.38; 95%CI 0.19 to 0.56; p<0.001).

**Table 2.**
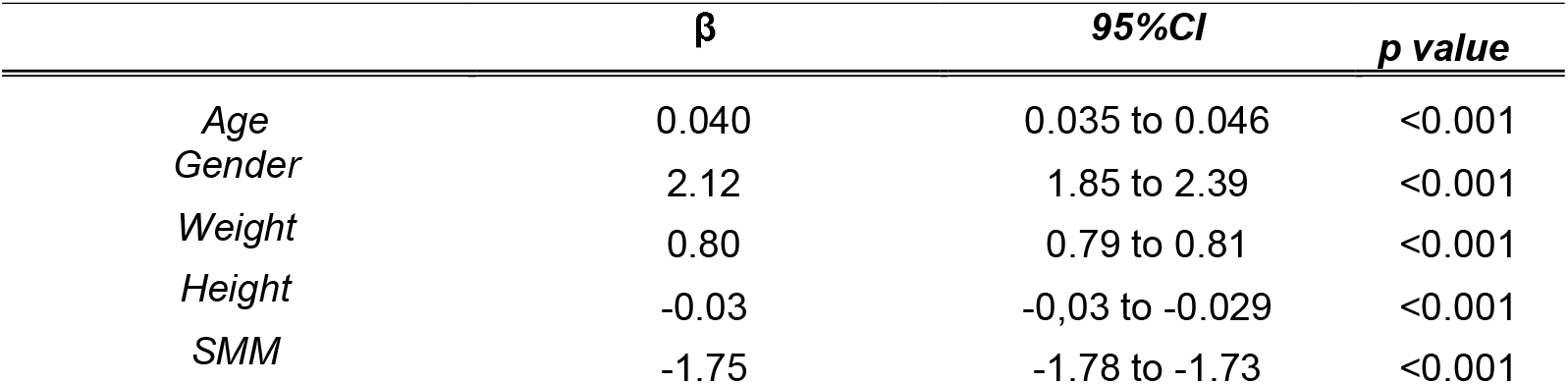
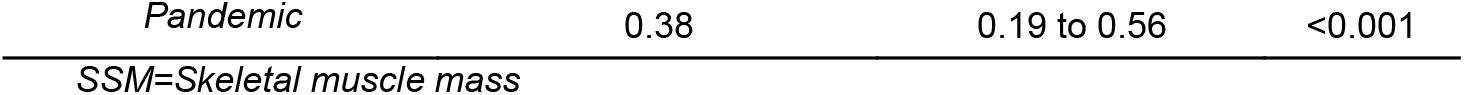
Linear multivariated regression analysis.

|  | **$\beta$** | **95%CI** | **p value** |
| --- | --- | --- | --- |
| Age | 0.040 | 0.035 to 0.046 | <0.001 |
| Gender | 2.12 | 1.85 to 2.39 | <0.001 |
| Weight | 0.80 | 0.79 to 0.81 | <0.001 |
| Height | -0.03 | -0,03 to -0.029 | <0.001 |
| SMM | -1.75 | -1.78 to -1.73 | <0.001 |
| *Pandemic* | 0.38 | 0.19 to 0.56 | <0.001 |

*SSM=Skeletal muscle mass*

For the second analysis 112 paired measurements were included. Table 3 demonstrate the pairwise comparison. Among BC variables only PBF increased in pandemic time compared to pré-pandemic. A small ES was again observed.

**Table 3.** Pairwise comparison of the 112 individuals with BIA in both time frames

| *N=112* | *Pre-pandemic* | *Pandemic* | *Cohen's d (95%CI)* | *p value* |
| --- | --- | --- | --- | --- |
| *Age (Years)* | 42.27±16.7 | 43.46±16.7 | 1.76 (1.48 to 2.05) | <0.001 |
| *BMI (kg/cm2)* | 25.59±4.48 | 25.82±4.58 | 0.10 (-0.07 to 0.29) | 0.264 |
| *FFM (kg)* | 57.53±12.35 | 57.51±11.91 | -0.01 (-0.19 to 0.17) | 0.948 |
| *SMM (kg)* | 32.27±7.52 | 32.2±7.27 | -0.03 (-0.21 to 0.15) | 0.722 |
| *BFM(Kg)* | 19.47±10.64 | 20.34±11.05 | 0.16 (-0.02 to 0.35) | 0.077 |
| *PBF (%)* | 24.59±10.34 | 25.58±9.89 | 0.23 (0.04 to 0.42) | 0.015 |
| *VFA (cm2)* | 86.9±53.68 | 90.5±52.69 | 0.16 (-0.02 to 0.34) | 0.093 |

*BMI=body mass index; BFM= body fat mass, FFM= free fat mass; SMM= skeletal muscle mass; PBF= percentage body fat; VFA= visceral fat area.*

## 4. DISCUSSION

To our knowledge this is the first populational research comparing BC before and after the SARS-CoV-2 pandemic.

Despite the non-differences in W, a less healthy BC profile characterized mainly by higher PBF (↑8.2%) and lower SSM (↓4,2%) was shown. After adjustment for some important health and BC determinants such as age, gender and SSM, the pandemic period of time was an independent PBF determinant. Likewise, paired sample showed a higher PBF on pandemic compared to pre-pandemic.

Since the beginning of the SARS-CoV-2 pandemic, many authors have been alerting for the risk of increasing unhealthy behaviors such as sedentarism, alcoholism, excessive energy intake, worsened diet composition and their consequences on metabolic health(10–12).

PBF is closely related to many cardiovascular risk factors, and even is a better predictor than the traditional and widely used BMI. Zeng at al.(13) shown a 3%, 5% and 3% risk increment on hypertension, dyslipidemia and hyperglycemia per 1% increase on PBF, respectively. Nagaya et al(14) showed that PBF was a better predictor of a worse lipid profile than BMI.

What would be the effect of such an increase in the PBF and cardiovascular risk factors in terms of hard clinical end points such as infarction, stroke and cardiovascular death?

This research has some limitations that worth to be highlighted. Frist, as a single center we cannot guarantee that the same profile will be observed among different population. Nevertheless, we hope that our results could encourage other researchers. Second, the pandemic brought more sick/unhealthy people to our clinic, mainly the COVID-19 survivors with long term symptoms. Although the pandemic period of time remained highly directly related to PBF even after adjusting for covariates, we cannot affirm that our results would be the same including comorbidities and physical activity behavior in the model. Again, this is a very interesting subject for future research.

In conclusion, our data show an increase in PBF and a decrease in SSM during the pandemic period of time compared to pre-pandemic. We reproduce here a part of the paper from Hall et al.(15) that brilliantly sums up our thoughts: *“…we are currently confronted with two pandemics occurring at the same time. The world will recover from the COVID-19 pandemic and so-called normal activities will resume. However, the physical inactivity/sedentary behavior pandemic will continue and, more troublingly, we may be at risk for this pandemic to worsen as a result of COVID-19. As a global society, we simply cannot let this happen*.*”*

It is comprehensible that, currently, health authorities’ efforts should be focus on SARS-CoV2 pandemic’s control such as vaccination and hospital facilities for critical cases. However, the potential chronic diseases burden, triggered by unhealthy behaviors must not be lost sight of. Public health policies should be implemented in this regard.

## Data Availability

The data that support the findings of this study are not openly available due to [reasons of sensitivity e.g. human data] and are available from the corresponding author upon reasonable request

## REFERENCES

1. Zheng C, Huang WY, Sheridan S, Sit CH-P, Chen X-K, Wong SH-S. COVID-19 Pandemic Brings a Sedentary Lifestyle in Young Adults: A Cross-Sectional and Longitudinal Study. Int J Environ Res Public Health 2020;17:.

2. Ferrante G, Camussi E, Piccinelli C, Senore C, Armaroli P, Ortale A, Garena F, Giordano L. Did social isolation during the SARS-CoV-2 epidemic have an impact on the lifestyles of citizens? Epidemiol Prev 2020;44:353–362.

3. Robillard R, Saad M, Edwards J, Solomonova E, Pennestri M-H, Daros A, Veissière SPL, Quilty L, Dion K, Nixon A, Phillips J, Bhatla R, Spilg E, Godbout R, Yazji B, Rushton C, Gifford WA, Gautam M, Boafo A, Swartz R, Kendzerska T. Social, financial and psychological stress during an emerging pandemic: observations from a population survey in the acute phase of COVID-19. BMJ Open 2020;10:e043805.

4. Henschel B, Gorczyca AM, Chomistek AK. Time Spent Sitting as an Independent Risk Factor for Cardiovascular Disease. Am J Lifestyle Med 2020;14:204–215.

5. Stamatakis E, Gale J, Bauman A, Ekelund U, Hamer M, Ding D. Sitting Time, Physical Activity, and Risk of Mortality in Adults. J Am Coll Cardiol 2019;73:2062–2072.

6. Williams MJ, Hunter GR, Kekes-Szabo T, Snyder S, Treuth MS. Regional fat distribution in women and risk of cardiovascular disease. Am J Clin Nutr 1997;65:855–860.

7. Vasan SK, Osmond C, Canoy D, Christodoulides C, Neville MJ, Di Gravio C, Fall CHD, Karpe F. Comparison of regional fat measurements by dual-energy X-ray absorptiometry and conventional anthropometry and their association with markers of diabetes and cardiovascular disease risk. Int J Obes (Lond) 2018;42:850–857.

8. Chen G-C, Arthur R, Iyengar NM, Kamensky V, Xue X, Wassertheil-Smoller S, Allison MA, Shadyab AH, Wild RA, Sun Y, Banack HR, Chai JC, Wactawski-Wende J, Manson JE, Stefanick ML, Dannenberg AJ, Rohan TE, Qi Q. Association between regional body fat and cardiovascular disease risk among postmenopausal women with normal body mass index. Eur Heart J 2019;40:2849–2855.

9. Kyle UG, Bosaeus I, De Lorenzo AD, Deurenberg P, Elia M, Gómez JM, Heitmann BL, Kent-Smith L, Melchior JC, Pirlich M, Scharfetter H, Schols AMWJ, Pichard C. Bioelectrical impedance analysis - Part I: Review of principles and methods. Clin Nutr 2004;23:1226–1243.

10. Clemmensen C, Petersen MB, Sørensen TIA. Will the COVID-19 pandemic worsen the obesity epidemic? Nat Rev Endocrinol 2020;16:469–470.

11. Chiwona-Karltun L, Amuakwa-Mensah F, Wamala-Larsson C, Amuakwa-Mensah S, Abu Hatab A, Made N, Taremwa NK, Melyoki L, Rutashobya LK, Madonsela T, Lourens M, Stone W, Bizoza AR. COVID-19: From health crises to food security anxiety and policy implications. Ambio 2021;50:794–811.

12. Calina D, Hartung T, Mardare I, Mitroi M, Poulas K, Tsatsakis A, Rogoveanu I, Docea AO. COVID-19 pandemic and alcohol consumption: Impacts and interconnections. Toxicol reports 2021;8:529–535.

13. Zeng Q, Dong S-Y, Sun X-N, Xie J, Cui Y. Percent body fat is a better predictor of cardiovascular risk factors than body mass index. Brazilian J Med Biol Res = Rev Bras Pesqui medicas e Biol 2012;45:591–600.

14. Nagaya T, Yoshida H, Takahashi H, Matsuda Y, Kawai M. Body mass index (weight/height2) or percentage body fat by bioelectrical impedance analysis: which variable better reflects serum lipid profile? Int J Obes Relat Metab Disord J Int Assoc Study Obes 1999;23:771–774.

15. Hall G, Laddu DR, Phillips SA, Lavie CJ, Arena R. A tale of two pandemics: How will COVID-19 and global trends in physical inactivity and sedentary behavior affect one another? Prog Cardiovasc Dis 2021;

